# Limited Research Investigating the Value of MRI in Predicting Future Cognitive Morbidity in Survivors of Paediatric Brain Tumours: A Call to Action for Clinical Neuroimaging Researchers

**DOI:** 10.1101/2024.01.12.24301212

**Authors:** Daniel Griffiths-King, Christopher Delivett, Andrew Peet, Jane Waite, Jan Novak

## Abstract

Survivors of pediatric brain tumour patients are at high risk of cognitive morbidity. There is clinical benefit in being able to reliably predict, at the individual patient level, whether a patient will experience these difficulties or not, the degree of impairment, and the domains affected. Whilst established risk factors exist, quantitative analysis of MRI could provide added predictive value towards this goal, above and beyond existing clinical risk models. The current systematic review aims to answer the question “Do MRI markers predict future cognitive functioning in pediatric brain tumour survivors?”. Studies of pediatric brain tumour patients which test the value of MRI variables in predicting later neuropsychological outcomes were searched up to March 2024. Only included were studies where MRI scans were acquired at an earlier timepoint and used to predict a child’s performance on cognitive tests at a later timepoint. Surprisingly few studies were identified by the systematic search process, but those that were identified specifically investigated MRI measures of cerebellar and white matter damage as features in predicting cognitive outcomes. Ultimately, the key finding of this review is that the current literature is limited. Those studies identified had small sample sizes and were rated as poor quality for the purposes of prediction. Therefore, current findings are at high risk of bias and thus the quality and conclusions are limited. Given the significant impact for this clinical population that predictive models would enable, the current review affirms the need for a ‘call to action’ for medical imaging researchers in pediatric neurooncology.

## Introduction

### Individual Outcomes for Childhood Brain Tumour Patients

Survival from cancer in childhood has seen great improvement in recent decades [1]. Consequently, there is an increasing population of adult survivors [1, 2], with approximately 1 in 530 young adults between the ages of 20 and 39 being a survivor of childhood cancer [3]. This is especially true in pediatric brain tumours, the most common solid tumours in children (roughly 20%) [4], where survival is now estimated at around 95% for cerebellar pilocytic astrocytoma, and 60-80% for medulloblastoma [5-8]. Thus, there is an ever increasing need to focus on ensuring quality of life for the future of these children.

Many children with brain tumours experience neurocognitive effects at some point in their disease course, resulting in dysfunction in domains of cognition, emotion, and behaviour. The estimated risk for children with brain tumours of having emotional, psychosocial, and attention problems are 15%, 12% and 12% respectively, according to a recent meta-analysis [9]. Even at 10-year survival, these patients still demonstrate neuropsychological and psychosocial impairment across multiple domains [10]. Recent, large-scale, longitudinal studies have suggested an increased risk of continuing neurocognitive decline for these patients, irrespective of treatment type [11]. Performance over time demonstrates an inability to acquire new skills and cognitive abilities at the same rate as healthy peers, rather than a loss of previously acquired abilities [12]. This may explain why these difficulties are likely to persist long-term and are non-transient. The number of post-cancer life-years is greater for pediatric rather than adult survivors, and these years include important milestones such as education and interpersonal relationship development [13]. Long-term difficulties could profoundly affect participation for these children, at home, school and later in the workplace, likely resulting in poorer long-term educational and employment outcomes [14, 15]. This represents a persistent burden for patients, families and healthcare systems [16]. Whilst survival must always be the utmost priority, research aimed at limiting cognitive morbidity in this group is now needed to ensure likelihood of reaching their potential, despite their illness [17].

Whilst disease and treatment will inevitably place all pediatric brain tumour patients at some level of risk for poor cognitive outcomes, knowing individualised risk, an estimate of the severity of difficulties and specific domains likely to be impacted, will influence clinical practice. There is significant variability in outcomes at the individual patient-level, but this is currently understudied [16]. Person-centred analytical approaches across a large longitudinal sample of paediatric brain tumour patients, show distinct classes / phenotypes with unique profiles in social, cognitive, and attentional difficulties over time [18], with similar subgroups identified in cross-sectional data [19]. Percentages of individuals scoring in the ‘impaired range’ was between 28-55% across domains in a recent longitudinal study, at around 6yrs post diagnosis [11] – highlighting, within a ‘cutoff’ driven framework of cognitive impairment, the presence of a classification task for identifying/labelling individual cases of impairment. Thus, there is scope for developing individualised models of risk and resilience, which hold predictive validity.

### Clinical Benefits from Prediction of Cognitive Outcomes

Prediction of individual-level neurocognitive outcomes would enable timely and tailored input from school and allied health services, promoting outcomes for these children, with limited healthcare resources being efficiently prioritised for those most at risk. It would also help healthcare professionals counsel and educate patients for these difficulties and help reduce uncertainty about the future for families. Individual models of risk would also impact treatment planning. In children where treatment of their brain tumour is more difficult, adjuvant therapy may include radiotherapy, which is known to have significant impact on a child’s neurodevelopment. This is due to brain injury from the primary and secondary effects of radiotherapy, especially in paediatrics where there is specific vulnerability (e.g., due to younger children not yet having reached peak myelin maturity) [20, 21]. Whilst developments in treatment have mitigated some neurocognitive toxicity (e.g. proton beam radiotherapy [22]), there is still need for clinicians to navigate treatment decisions in terms of risk to QoL based upon known disease and age related risk factors [20, 21]. More accurate prediction of individual-level risk of cognitive morbidity (even across domain and severity), would enable clinicians to further adapt and personalise treatment schedules with a greater focus on risk to quality-of-life whilst maintaining treatment efficacy [23]. Overall, there is clinical benefit for a range of patients in knowing individualised prediction of neurocognitive outcomes, and developing these methods for deployment to a clinical setting.

### Predicting Cognitive Outcomes

There are many established risk factors for poor long-term neuropsychological outcome that need to be understood to provide a comprehensive risk profile at the individual child level [24]. Recent neurodevelopmental models based on known risk factors have been proposed to explain outcomes for brain tumour survivors, specifically in medulloblastoma [25-27], taking into consideration the complex disease-, treatment- and host-related factors that may influence these outcomes. Many aspects can result in neurodevelopmental insults to the developing brain which may explain and underpin these neurobehavioral morbidities [28, 29] and thus are significant risk factors for these poor outcomes [24]. These range from physical factors such as treatment effects (i.e. resection and/or adjuvant therapy [20, 30]) and individual differences (e.g. age at diagnosis [30], cognitive reserve), but also psychological factors (i.e. Early Childhood Adversity, threat exposure) and environmental factors (i.e. Socioeconomic Status (SES) and social support) [28]. See [24] for a model of cognitive risk in pediatric brain tumour survivors. Essentially, neurocognitive outcomes are complex and are dependent on several interacting factors [13].

Risk-based and exposure-related guidelines and models have been developed to manage these neurocognitive late-effects of pediatric brain tumours [24, 31]. Neurobehavioral morbidities are predicted by clinical variables such as radiotherapy, chemotherapy, neurosurgery, and parental education but less-so age at diagnosis, gender, or time since diagnosis [13, 14, 20, 32-35]. A number of these complex risk factors can be either difficult to measure or qualitative in their assessment and therefore can inform decisions but do not make individual predictions. The Neurological Predictor Scale (NPS) was designed as an ordinal scale to quantitatively capture the cumulative effect of several risk factors on outcomes, and somewhat predicts IQ, adaptive functioning and processing speed and working memory, at both short- and long-term follow-up [34, 36-39]. This cumulative measure captures unique variance, above and beyond the individual predictors.

### MRI as a Novel Predictor of Outcomes

This systematic review posits that magnetic resonance imaging (MRI) measures are likely to be a good proxy of the burden of brain tumours and their treatment thus, are likely to be predictive of cognitive impairment at the individual patient-level. Qualitative reporting of MRI does predict outcomes, with brainstem invasion, midline location of the tumour, and tumour type predicting post-operative cerebellar mutism syndrome, a (typically) transient, neurological morbidity seen in this population [40]. Quantitative alterations to the brain’s structure and function, specifically microstructural changes to the white matter (WM) of the brain, during the developmental period, could be the common neuroanatomical substrate of poor neurocognitive outcomes [25, 27]. See [41] and [23] for a review of MRI in pediatric brain tumours. Recent successes and interest in using MRI to predict neurodevelopmental outcomes in premature infants [42], or even decline in neurocognitive functioning in older adults [43] highlights the potential opportunities offered by MRI. There is also a relative abundance of MRI data in these patients, acquired as part of standard of care and most research protocols. Therefore, MRI is likely to provide highly relevant features which provide ‘added-value’ in predicting outcomes beyond clinical risk factors alone.

A key consideration, however, is the timing of the MRI used to predict these outcomes. The MRI used for prediction needs to be early enough in the disease course and non-contemporaneous from the later, outcome of interest. There is currently no consensus on the optimal timing of MRI with which to make such a prediction. This systematic review specifically investigates existing literature using MRI scans, taken at any point in the disease course, to predict non-contemporaneous, later neuropsychological outcomes in survivors of pediatric brain tumours.

Whilst there is existing literature of existing established clinical predictors of cognitive late effects in this population, this review aims to assess studies using MRI as a predictive modality, with the goal of assessing whether quantitative analysis of MRI provides ‘added-value’ in these risk models.

## Method

We conducted this systematic review in accordance with Preferred Reporting Items for Systematic Review and Meta-Analysis (PRISMA) guidelines [44], an overview of which is reported in Figure 1. Initially, a limited search of the Web of Science database was undertaken in June 2022 for the purpose of refining the search terms. Due to the wide-ranging classifications of central nervous system (CNS) tumours, as well as generic tumour-focussed terms, we also included terms pertaining to the most common paediatric histological diagnoses accounting for 85% of total incidence rates (Central Brain Tumour Registry of United States, 2014-2018 [45]). Search terms can be found in supplemental materials. Based on our initial search, we pre-registered our review protocol through the International Prospective Register of Systematic Reviews (PROSPERO) database (registration number CRD42022343161).

**Figure 1.**
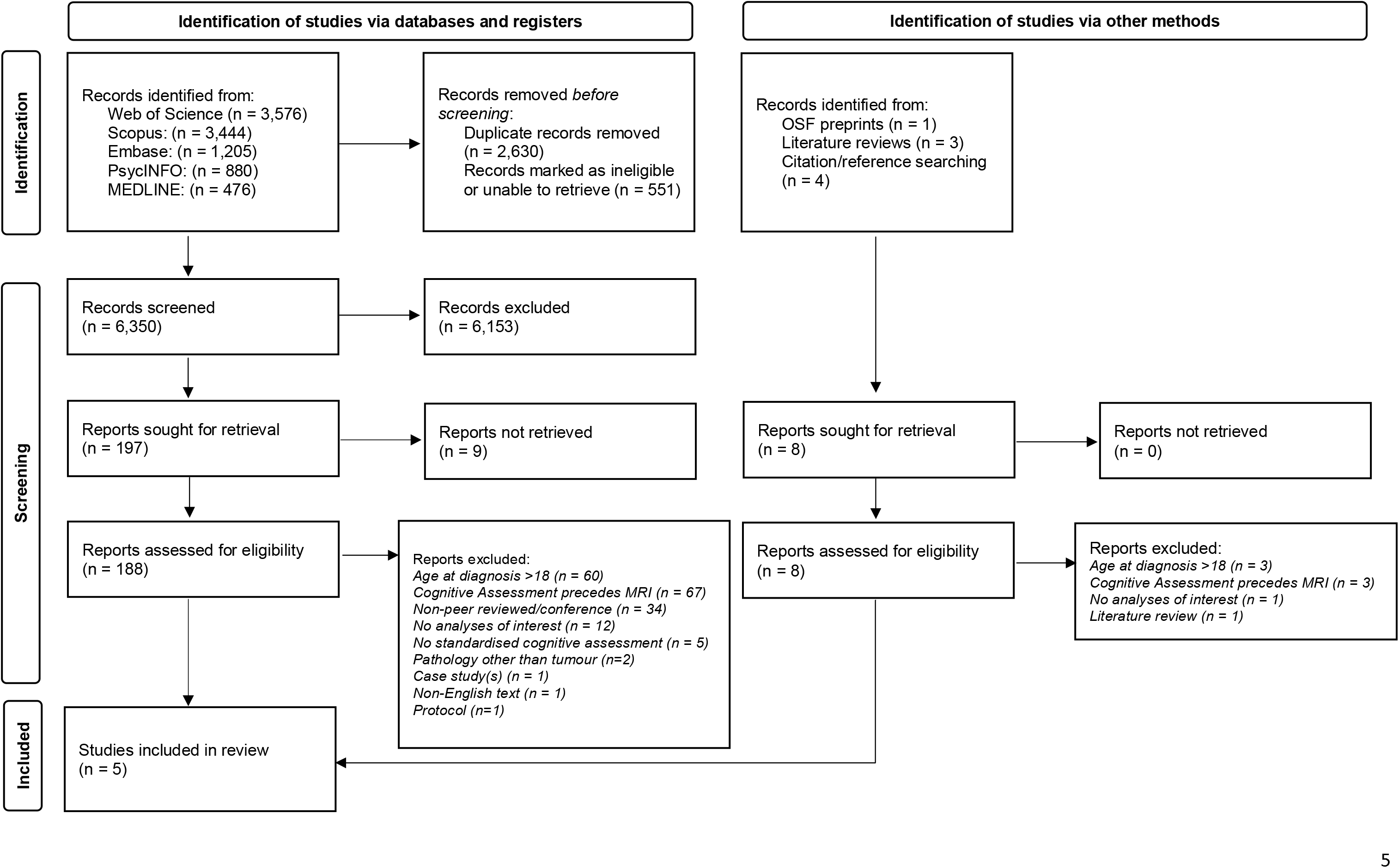
PRISMA flow diagram (based on March 2024 Search)

A comprehensive search of Embase, MEDLINE, PsycINFO, Scopus and Web of Science was conducted in July 2022 using the designed search, resulting in 8,632 records. Searches were rerun and results updated in March 2024, resulting in an additional 899 records. Alterations were made to the search terms for each database to account for differing Boolean operators (see Supplemental Materials). Additionally, we also searched the Open Science Framework (OSF) preprints archive for relevant articles that had not otherwise appeared as published texts in our main search. We included any longitudinal study concerning patients diagnosed with a brain tumour before the age of 18, who had MRI data that clearly preceded an age-appropriate, standardised test of cognitive ability (e.g., intellectual ability assessed with WISC-V [46]). Central to our main research aim, we included those studies that explicitly reported an association between future cognitive outcomes based on prior MRI. Meta-analyses and literature reviews that did not report new data were excluded, however, reference lists of relevant papers were searched for additional studies of interest. Search results were not restricted by publication date but were limited to those written in English. In addition to our pre-registered exclusion criteria, we also excluded patients with neurofibromatosis, tuberous sclerosis, or acute lymphoblastic leukaemia as these were considered significant confounds for predicting cognitive outcomes. We also excluded non-peer reviewed articles, such as conference abstracts and theses. Inclusion/exclusion criteria are further detailed in Table 1.

**Table 1.**
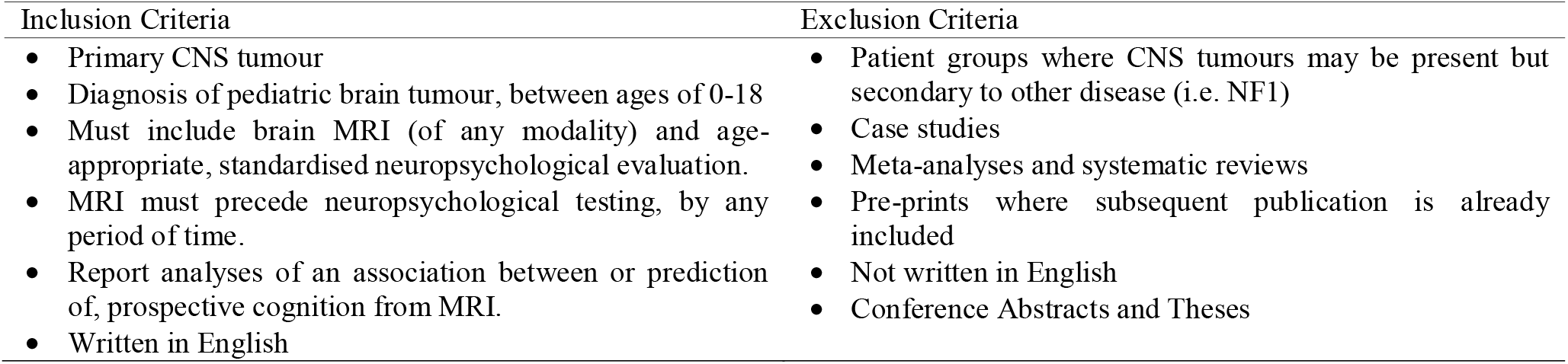
Inclusion and exclusion criteria for identifying publications for the systematic review.

Identified records were first imported into MS Excel and duplicates removed. Following a short pilot, two independent reviewers (CD + DGK) screened the titles and abstracts of all the identified papers against the inclusion criteria. Full texts of suitable papers were subsequently retrieved and screened by both reviewers for final inclusion in the review. For completeness, the reference lists and citations of those papers marked for inclusion were reviewed for additional studies that may have been missed. At each stage of the process, disagreements were discussed until consensus was met. Per our pre-registration, data extraction was completed by one reviewer (DGK), whilst a second reviewer evaluated data extraction of all papers for correctness (JN). The data extraction tool was initially developed for this research protocol and was later refined based upon the findings of the search results. This was not based on an existing tool, and items were selected based on discussion within the research team. Data from each study included: (1) year of publication, (2) study aims and/or hypotheses, (3) study location (i.e., country, recruiting hospital), (4) number of patients, (5) patient characteristics (i.e., years recruited, diagnoses, treatments, cognitive outcomes, age at diagnosis/MRI/neuropsychological evaluation), and (6) statistical analyses.

We had initially registered our intention to assess the validity of the included studies using the Transparent Reporting of a multivariate prediction model for Individual Prognosis or Diagnosis (TRIPOD) guidelines; however, this was deemed unsuitable given that none of the studies reported using predictive modelling in their approach. Instead, studies were reviewed (by DGK) using the Reporting Recommendations for Tumor Marker Prognostic Studies (REMARK) checklist [47, 48], a checklist for assessing reporting quality specific to the domain of oncology. Whilst designed for marker/assay testing, the domain relevance and prognostic nature renders this a relevant tool. We considered the MRI measures as the ‘marker’ under investigation and the neuropsychological assessment as ‘endpoints’ for the purposes of the checklist.

## Results

After reviewing titles and abstracts, 197 records were selected (see Fig1), and 188 full-text articles were assessed for eligibility. Of those, five studies were included. Manual reference and citation checking of these selected articles (and identified literature reviews deemed to be relevant), identified no additional studies. Detailed information about the included studies can be found in Table 2.

**Table 2.**
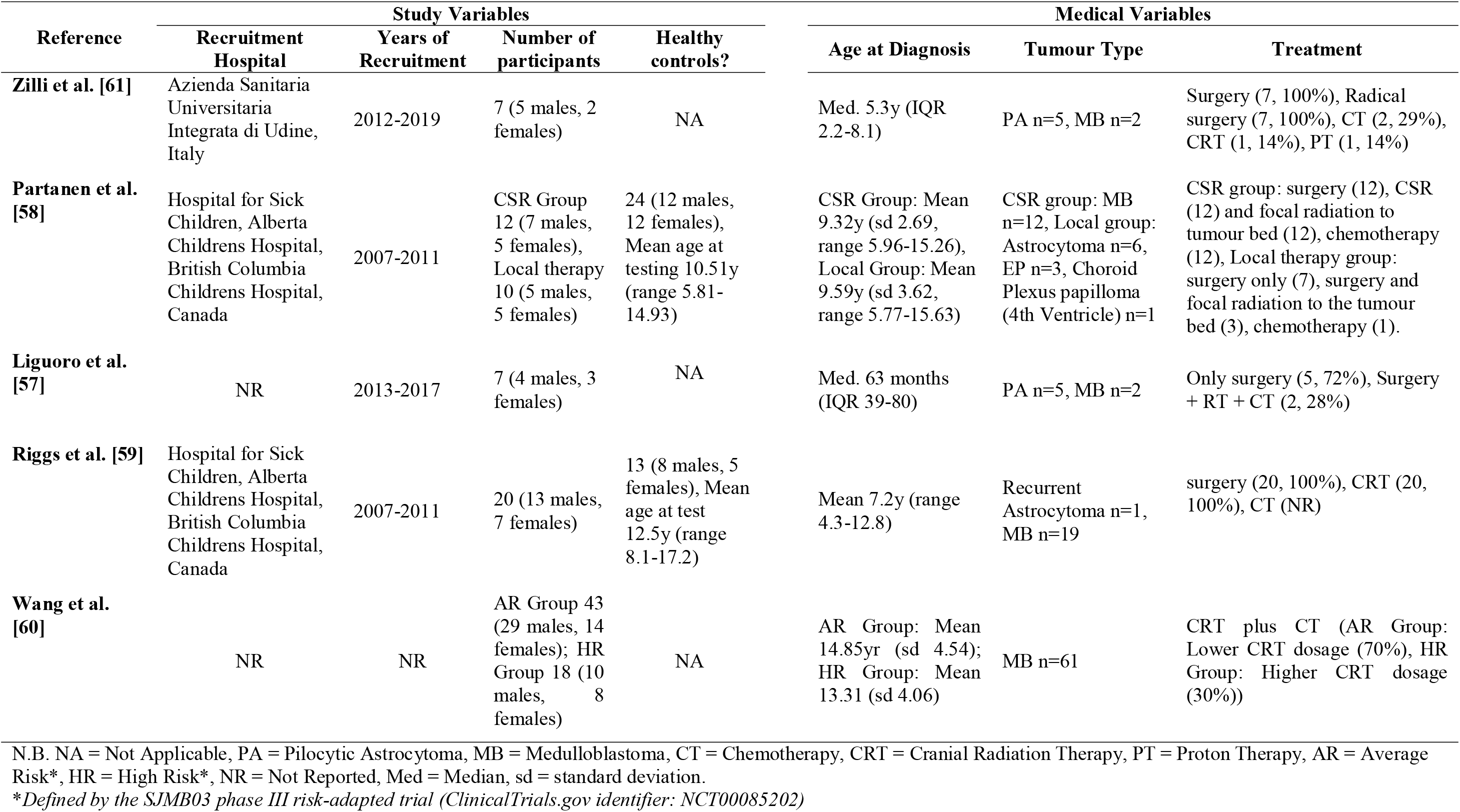
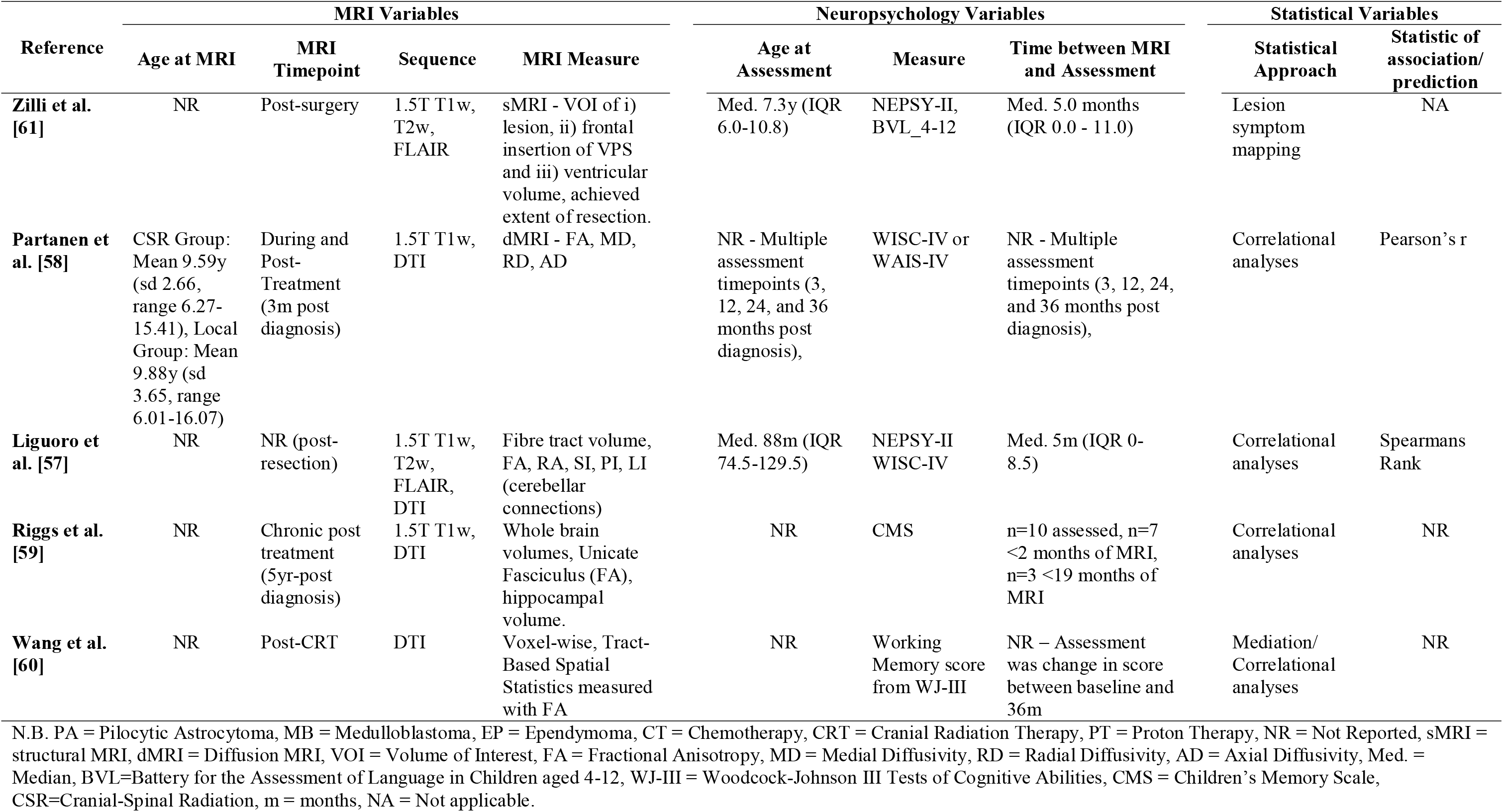
Details of included studies

### Study characteristics and reporting quality

Many studies were excluded because the MRI did not precede neuropsychological assessment (for instance because of the matched timepoints of neuroimaging acquisition and test assessment), thus not defining them as ‘predictive’ studies. In a small number of cases, the text was ambiguous to the order of testing (i.e., [49-54]) but did not refer to prediction or other details suggestive of the order, and thus were not included. For other studies, data including MRI which preceded a later neurocognitive assessment existed, due to the inclusion of multiple timepoints, however it was ambiguous in the analyses of interest as to the time points being referred too and so these studies were not included [55, 56].

For the selected studies, sample sizes were small and ranged between n=7 and n=61; altogether (notwithstanding dataset overlap) only n=118 pediatric brain tumour patients and n=37 healthy controls were included across the reviewed studies. The most common tumour type across studies was medulloblastoma (n=96), then astrocytoma (n=17) with relatively few ependymoma and choroid plexus papilloma (n= 3 & 1 respectively). Age at diagnosis across the studies ranged from 2.2 years to 15.6 (based on ranges and inter quartile range (IQR)). All studies selected associative statistical approaches (i.e., correlational analyses), with one also adopting a mediation approach.

Using the REMARK checklist, studies were assessed against each reporting item (*Item 1-20*), and here we report items where reporting was limited across the studies (i.e. one or less studies reported the item). No studies gave a rationale for sample size (Item 9, 0/5 studies), likely due to the limited samples in each study, however it was unclear as to whether these were the entirety of eligible patients within the given timeframe (as only 1 study gave a full accounting of the flow of patients in the study, *Item 12*, 2/5 studies) In terms of “Analysis and presentation”, studies performed poorly for a number of items (*Item 15, 16, 18*, all 1/5 studies, and *Item 17 0/5* with no studies completing the item to be reported). Firstly, only one study presented an effect size for the predictive analysis (*Item 15*, 1/5 studies). Further, included studies did not conduct analysis of added value, including the MRI marker and ‘standard prognostic variables’ which are established (*Item* 17, 0/5 studies) nor sensitivity analysis/validation although one study confirmed statistical/theoretical assumptions (*Item 18*, 1/5 studies).

### White Matter (WM) predictors

Of the studies assessed, three utilised diffusion tensor imaging (DTI) to image white matter as a predictor of outcomes. Liguoro et al. measured the fractional anisotropy (FA) and volumetry of spinocerebellar (SC), dentorubrothalamocortical (DRTC) and corticopontocerebellar tracts (frontopontocerebellar (FPC), parieto-pontocerebellar (PPC), occipitopontocerebellar (OPC), and temporo-pontocerebellar (TPC)) [57]. Significant relationships were found between tracts relevant to cerebellar connectivity, and the Developmental Neuropsychology Assessment (NEPSY) and full-scale IQ (FSIQ) measured approximately 5 months later [57]. Specifically, FSIQ correlated significantly with spherical and planar indices of the right PPC (r=-1, p=0.017 and r=0.886, p=0.033), with increases to planar index and decreases in spherical index associated with IQ [57]. Liguoro et al. also found significant correlations between specific fibre tract characteristics and tasks measuring attention, memory, sensorimotor, social perception, and visuospatial processing domains. However, only visuospatial processing showed convergent validity with significant correlations across two different tasks measuring this same domain [57]. In this study, the bilateral PPC and SC tracts were most commonly correlated with the neuropsychology tasks [57].

Partanen et al., used MRI from the treatment period 3 months after diagnosis (including during and after treatment) to predict change in intellectual functioning over a 36 month period after diagnosis. A significant reduction in FSIQ over time was found but this was not related to diffusion measures (FA, mean diffusivity (MD), radial diffusivity (RD) and axial diffusivity (AD)) for the cortical spinal tract (CST), inferior fronto-occipital fasciculus (IFOF), inferior longitudinal fasciculus (ILF), optic radiations (OR), and uncinate fasciculus (UF) [58]. Partanen et al. did however show that declines over time in processing speed index, observed only in a subgroup of patients experiencing local therapy (i.e., focal radiation) versus cranial spinal radiation, was predicted by baseline anisotropy in left inferior fronto-occipital fascicle (IFOF), with lower FA being related to greater decline [58]. Neither patient groups showed a difference in the left IFOF for diffusions measures compared to controls.

Riggs et. al. [59] utilised chronically acquired MRI (approximately 5 years post diagnosis) investigated correlations between whole brain WM volume, FA of both the left and right UF and the general memory index of the Childhood Memory Scale (CMS) (in a subset of n=10 patients, outcomes measured between 2 and 19 months after MRI). Only FA of the left UF was significantly related to memory (R=.64, p=.045), not the right uncinate fasciculus or total WM volume (as measured by structural MRI), with increased FA related to increases in memory performance. The volume of the PPC tract also positively correlated with memory performance (R=.71, p=.045) in Parten’s study [58].

Wang et al. [60] used a high-dimensional mediation model to estimate microstructural damage to brain WM that mediates the negative treatment effects craniospinal radiation has on declining working memory outcomes over a 36month period. Post-treatment DTI was used to estimate FA tract-based spatial statistics (TBSS) across tracts within a white-matter atlas. Larger FA was associated with better working memory outcome, across multiple WM tracts including the cerebral peduncle, corpus callosum, splenium and corona radiata. Specifically, Wang et al [60] found that there was a significant negative mediation effect of the WM microstructure between radiation treatment (average risk / lower dose vs high risk / higher dose) and the change in working memory over 36 months. This study therefore demonstrated the causal effect of radiation-related damage to white matter predicting long-term working memory in these children, accounting for around 43% of the overall impact of treatment on long-term working memory decline.

Strength of correlational relationship between indices of white matter integrity and neuropsychological outcome were large (according to Cohen’s criteria) ranging from |r|=.64 - |r|=1. However, the very limited sample sizes (n=10 & n=7) from which these were drawn gives reason for concern over the interpretation of these estimates.

### Grey Matter (GM) predictors

In Riggs et. al., no correlation was found between total GM volume or left hippocampus with the general memory index of the CMS, but the right hippocampal volume, measured at a chronic timepoint, showed significant positive correlation with memory outcomes 2-19 months after MRI (R=.71, p=.02) [59]. It is important to note in this study, that the right hippocampus, rather than the GM volume and left hippocampus, was significantly smaller in the patient group compared to healthy controls.

### Lesion predictors

Zilli et. al. [61] used a lesion-symptom mapping approach, to investigate the overlap of lesions in children with versus without psychological impairment. The lesions investigated where tumour lesions, frontal insertion of ventriculoperitoneal drainages and ventricular volumes, as drawn on the T1w MRI. They found the greatest tumour lesion overlap and therefore greatest damage was found in median cerebellar, specifically paravermal and vermal regions. Regions of interest for the lateral ventricles also overlapped in impaired children, suggesting hydrocephalus as additional cause of future impairment.

### Meta-analysis

Despite registering our intention, reviewed studies were not of sufficient quality to conduct any form of meta-analysis due to varying measurement strategies, gaps in reported descriptive variables, and low power.

## Discussion

### Review of the State of Research

The current systematic review aimed to investigate existing literature using MRI to predict later, and non-contemporaneous neuropsychological outcomes in children with brain tumours. No studies reviewed here set out the rationale for and/or aimed to predict future outcomes using model development and validation approaches. The lack of scientific attention given to this topic is surprising given the dearth of literature advocating for such research. The papers identified and reviewed here, did in fact conduct analyses to this effect, but only due to the fortuitous nature of the selected timepoints, and intervals between the activities of MRI scanning and neuropsychological testing. Despite an extensive search strategy, evidence with which to answer the current research question was extremely limited, with the major finding being a severe lack of studies in this area. The primary result of the review must therefore be viewed as a need for further research in this very important research area, with study designs that directly tackle the need for outcome prediction in these cohorts.

The reason for this limited number of research studies is unclear. Whilst our systematic search strategy was extensive, there were also difficulties in identifying papers due to poor reporting practices. For instance, in some studies, the timing of MRI in relation to assessment was ambiguous or unclear [49-52, 56]. Another potential cause of limited research could be previous focus on survival, where increasing survival rates are now placing a greater need for research on late effects. It is important to also consider that neurocognitive effects are also only one of many potential late effects experienced by this population [62, 63].

Quantity of research in this area may also be impacted by the availability of clinical data with which to carry out this research. For children with pediatric brain tumours, there is an abundance of clinical MRI, with medical imaging required for vital for tumour detection and diagnosis, surgical and radiotherapy planning, and monitoring of treatment response and recurrence of disease. But this is not necessarily echoed in access to neurocognitive assessment; testing is performed based upon clinical need or clinical trial protocol. This potentially limits available retrospective datasets. This data also comes from a heterogeneous cohort, with these children facing heterogenous brain injuries as a result of their disease and treatment. Identifying homogenous patient groups inevitably results in the smaller sample sizes seen in the current studies. Overall, these factors are liable to impact the quantity of research studies in this field.

Whilst number of studies was limited, the quality of existing studies was also a significant limiting factor for the usefulness of research studies in this area. In the reporting quality assessment (using the REMARK checklist [47, 48]) identified studies did not meet important criteria for development of prognostic markers. Specifically, studies failed to conduct additional analyses necessary for this development, such as sensitivity or ‘added-value’ analyses – although this was likely due to limited sample sizes, therefore lacking statistical power necessary for these additional analyses. Studies are typically involving “retrospective, monocentric study investigating a pediatric disease with low annual incidence” [61] however future work will require larger sample sizes than those of the studies presented here. To note, the checklist also comments on several items pertaining to model building and multivariate analyses, which were not conducted in the current studies.

Overall, the findings from the reviewed research are limited – they have limited sample sizes and are rated as low quality in terms of prediction studies. Without proper validation and replication, the quality and impact of any conclusions must be viewed as limited and/or potentially spurious. However, the findings are briefly discussed here in terms of wider literature. This should be seen in the context of guiding hypothesis-driven future research and/or promoting future validation and/or replication of these findings.

### Summary of Findings of Reviewed Studies

#### Cerebellar Damage

Given common posterior fossa presentation in pediatric brain tumours, it is unsurprising that multiple studies in this review a-priori selected regions of interest within cerebellar structures and related fibre projections from this anatomical structure. Damage to these circuits predicted outcomes [57], with lesions to the median cerebellar regions common in cognitively impaired patients [61]. Studies of contemporaneous MRI and neuropsychology measures have found similar. Horská and colleagues [64] found a decrease in vermis volumes over a 6-month period were significantly related to radiation dose, and final volume after this period related to neuropsychological measures of motor speed. Significant recent evidence suggests that the posterior cerebellar lobes are key in maintain cognitive performance [65], and animal models suggest that intact cerebellar activity is required to enable typical developmental trajectories of cognitive abilities (in mice) [66]. Essentially, the cerebellum plays as an integral node in many distributed neural circuits that underpin multiple cognitive functions [67, 68]. Radical cerebellar resection has also been associated with extensive WM microstructure changes across the brain [69]. Overall, it is unsurprising that damage to cerebellar regions (through injury and treatment effects) may lead to and/or predict multiple cognitive morbidities.

#### WM damage

Riggs et al [59] argued that global measures of WM may be indicative of general injury and thus correlate well with general ability, however, integrity of discrete tracts (such as the UF) may be a better predictor of specific cognitive abilities – in this case memory. Previous reviews of cross-sectional research suggests a model where disorganised WM microstructure is related to poorer cognitive abilities, especially processing speed and memory deficits [23], by indicting that this ‘damage-related impairment’ is established early, and therefore WM microstructure is a potential biomarker to predict later impairment. Both preclinical and patient studies suggest a loss of both GM and WM volume, and failure of normal WM gain in pediatric brain tumour survivors [16]. There are multiple mechanisms of WM damage; hydrocephalous having direct neurotoxic effects on periventricular WM due to decreased perfusion and oedema [70] or intragenic effect of adjunct therapy (chemo and radiotherapy) as measurable by reduced volume and alterations to microstructural properties and failure of expected WM development [51, 71]. WM damage is likely non-transient; for instance, in non-irradiated patients 15 years after diagnosis, FA measures are reduced and are associated with impaired cognitive flexibility [72]. What is apparent is that, across treatment and disease effects, the subsequent WM injury is relevant to poor outcomes, across emotion, cognition, and behaviour [14, 51, 73].

### Specific issues with current research

#### Limited longitudinal studies

The biggest limitation of the current research field is the lack of longitudinal research answering this research question. Whilst there have been multiple studies understanding the contemporaneous neuroanatomical substrates of poor neuropsychological outcomes in pediatric brain tumours, from acutely post treatment to very long-term survivors, these have not translated into similarly large body of work understanding long-term risk (as highlighted by our findings) of cognitive morbidity. Further longitudinal research is needed to assess whether the contemporaneous neuroanatomical substrates of long-term impairment are in fact predictive in the context of longitudinal studies.

These longitudinal studies would also provide an opportunity to disentangle the developmental and age-related effects on this prediction-task. For several of the measures highlighted in this review (FA/MD etc.) there are known developmental trajectories [74] which will necessarily interact with disease-related changes. There is also likely to be unique effects of brain insult, across tumour growth, and treatment related injury at different ages, resulting in varying levels of long-term impairments [75]. The field will need to rely upon longitudinal studies (with sufficient sample size/statistical power) that can sufficiently disentangle these interactions.

#### Study Variables: Timing

The current systematic review includes studies that use MRI from any point in the disease course. The timepoint of the MRI used for the purposes of prediction in the reviewed studies were most commonly post some form of treatment (surgical or post radiation therapy, e.g. [60] and [59] respectiverly) other than 57 which included MRI during treatment. Given the limited research available, this was done to assess the entire literature, but results from different timepoints in the disease to conduct prediction will undoubtedly have varying interpretations. For instance, post-treatment MRI may identify insult-related factors which are related to later decline – as demonstrated by the study by Wang and colleagues [60]. Pre-treatment MRI may allow us to identify specific vulnerabilities to the longer-term neurocognitive effects – for instance Zheng and colleagues [76] propose that functional network plasticity pre-treatment may mediate the impact of surgery on later cognitive ability. However, any MRI timepoint is likely to capture a mixture of these two influences, vulnerability, and insult factors, which may contribute towards prediction.

Overall, there is no consensus on the optimal timing of MR imaging to use for predictive purposes. Selection of which MRI is likely to be most predictive (in terms of reliability, accuracy etc.) will not be trivial for future research. We propose that for future research, selection of MRI timepoints with which to test predictive validity should be guided by two principals – a) clinical need, and b) evidence-based theoretical grounds. For instance, in terms of clinical need, if the most useful purpose of these models is to aid/supplement treatment management decisions, then an early, pre-treatment MRI will be necessary. In terms of guiding MRI timing based upon existing evidence a strong example of this is the study by Wang and colleagues [60] which suggests there is a treatment related ‘injury’ which mediates radiotherapy—related working memory impairment, suggesting post treatment MRI would have benefit. Timing is an even greater consideration in this patient group compared to adult brain tumour patients due to the likely interaction also with ongoing brain development over time for these patients.

These children undergo MRI scanning at a number of timepoints in their disease course (e.g. diagnostic imaging, pre- and post-surgical evaluation, progression monitoring etc), and so there is significant data for potential retrospective studies to investigate effect of MRI timing on prediction. Direct comparisons between models using MRI from different timepoints will be meaningful to understand variation on predictive validity over time, and further inform designs for prospective predictive studies.

Timing of neuropsychological assessment is also not to be overlooked. To develop predictive models, a given endpoint will need to be set (for instance a given number of years post diagnosis). Overall neurocognitive trajectory is “idiosyncratic” over time, with longitudinal studies suggesting an injury-related early impact, followed by a decline or failure to meet the normal developmental trajectory and potential long-term plateauing [11, 18]. Therefore, the endpoint of interest, may also inform the timing of MRI which may be more predictive of longer-term outcomes.

#### Added Value of MRI

A major limitation of the current state of the research literature in this field is that the added value of MRI in prediction has not been established, above and beyond existing approaches. No reviewed studies assessed existing risk factors in a multivariate analysis to test the relative contributions, and therefore added value, of early MRI in predicting future neurobehavioral morbidities. However, Partanen et. al. reported that none of the medical or treatment variables that they tested predicted change in IQ scores over time [58]. This is despite these medical variables (Neurological Predictor Scale and presence of cerebellar mutism syndrome) predicting acute/contemporaneous neuropsychology outcomes, and MRI-derived measures of baseline WM injury being significantly related with outcomes [58]. This is limited evidence to support the incremental validity of MRI as a predictor of long-term outcomes. Future studies should ensure to test for unique and additional predictive power offered by quantitative MRI variables

#### Study Variables: Approach to ROIs

Across the studies reviewed here two conducted analyses in regions-of-interest (ROIs) directly related to sites of brain insult in these patients [57, 58], one in ROIs related to the cognitive comorbidity under investigation [59], and only one investigating characteristics of the lesion itself [61]. This does not consider how the wider brain network may be influenced by the brain tumour, and this information may explain/predict additional variance in outcomes. For instance, in paediatric neurological disorders/syndromes, differences in brain morphometry or connectivity have been found beyond the site of pathology (i.e. paediatric epilepsy [77]) or in the absence of frank pathology (i.e. mild paediatric TBI [78], MRI-negative epilepsy [79]). Disconnectome symptom mapping, shows that non-homologous lesions to the same brain network can generate the same cognitive sequalae in terms of deficits. [80]. Many compensatory and ‘rerouting’ models of functional brain activity post injury suggest that regions beyond the focal lesion may explain some sparing of cognitive abilities (another important factor in predicting endpoint neurobehavioral morbidities). These findings all show that disparate, diffuse, and non-lesioned changes to the brain, including tissue which may be typically thought of as ‘spared’, could also explain variance in neurobehavioral morbidities. Connectivity approaches to MRI have shown utility in contemporaneous measurements of MRI and neuropsychology [81, 82]. These neurobiological effects of injury beyond the focal lesion may provide further prognostic information towards the aim of a predictive model, however, to test a greater number of regions larger sample sizes will be necessary to accurately estimate statistical models across many more ROIs. This highlights one of the key challenges for future studies in this field being data collection.

### Recommendations for future research

Beyond the apparent requirement for more research studies in this field, there are specific recommendations that should guide future endeavours. In many cases, due to the rarity of disease, multinational and multicentre analyses will be needed to achieve the sample sizes necessary to definitively address some of the issues in this review. To do so, a level of harmonisation amongst research groups in terms of data collection is necessary to facilitate combining of cohorts. For instance, adhering to similar imaging protocols (following guidelines for advanced MRI in pediatric CNS such as those proposed by the European Society of Pediatric Oncology (SIOPE) [83]). Harmonisation will not only allow integration of multiple datasets, but potentially reduce biases in measurements caused by differences in MRI acquisition protocols.

Additionally, harmonisation of neurocognitive assessment will also facilitate data aggregation. In the absence of a common outcome measure for these children (for instance a common data elements set as proposed by the National Institute of Neurological Disorders and Stroke (NINDS) for other neurological disease [84]) broad composite measures should be used that can capture multiple aspects of the neurocognitive morbidity experienced by these patients (for example the Wechsler Intelligence Scale for Children [46] or the NIH Toolbox Cognition Battery [85]. Neurocognitive assessment protocols are being developed for specific tumour groups (for instance in childhood ependymomas [86]) which will provide practical approaches to “strongly support the routine incorporation of neuropsychology assessments as key outcomes” to “facilitate successful global collaborations” [87]. These should be adopted wherever possible.

To facilitate future reviews such as this, and more importantly meta-analyses of said future research, greater reporting expectations should be placed on researchers – given the current review highlighted this to be a key weakness in existing research. Emphasis should be on using reporting guidelines, and quality assessment checklists (such as the REMARK checklist used in the current review [47], or the tools provided by the EQUATOR (Enhancing the QUAlity and Transparency Of health Research) Network [88] such as the TRIPOD tool [89]). Transparent and full reporting will allow better assessment of the literature across the field. These recommendations will help facilitate the important goals of this research, hopefully leading to greater clinical impact.

### Limitations of Review

It should be noted that, despite an extensive search, no study explicitly investigated the research question of whether MRI could be used for long term prediction of neurocognitive outcomes in pediatric brain tumour patients. Described studies were reviewed here due to non-primary analyses which fulfilled inclusion criteria, and therefore it may be the case that other studies with such analyses may have been missed in the review process (for instance if these secondary analyses were not mentioned in the abstract). To address this, we erred on the side of caution in reviewing abstracts, using full-text review as a method to identify these relevant secondary analyses.

## Conclusion

As early as 2008, it was proposed that to truly balance the aggressiveness of treatment for childhood CNS tumours, against the relative quality of life due to cognitive impairment, an important factor is knowing the likelihood of any one individual experiencing neurocognitive impairment [13]. This individualised risk is key in purported models of monitoring and managing of neurocognitive functioning in children with brain tumours [90]. There has also been significant work in the field of pediatric brain tumours proposing developmental cognitive neuroscience models of late effects in survivors [13, 23, 91, 92]. These models, built on contemporaneous measures of cognition and brain development, alongside cross-sectional data, are inherently limited. Knowing individualised risk of long-term cognitive morbidity ahead of time would have significant clinical impact; to inform clinical management, prioritise resources/support, and reduce uncertainty for families. Overall, there exists plenty clinical reasoning to prompt scientific enthusiasm and attention for this topic.

However, despite these early calls for prediction, and models with which to guide these predictive studies, this systematic review highlights that the number of truly predictive studies (requiring a period between predictive features and long-term outcomes) is still limited. In conclusion, Given the increased number of adult survivors of childhood brain tumours, the poorer long-term cognitive, educational and employment outcomes [10, 14, 15] and the significant burden this represents to patients, families and healthcare, work now needs to be completed to integrate predictive data into these models, which will expand their explanatory value and utility to clinical practice. This will be an important next step in delivering further clinical impact for this patient group.

Given the great potential that MRI provides in investigating neurobiological effects of disease and treatment at the individual-level, the plethora of multimodal imaging available in these clinical populations and finally the positive clinical benefit this could offer, there is exciting opportunities for this type of research.

## Data Availability

Data extracted from included studies is publicly available and found (in its entirety) in Table 2. Lists of reviewed studied, at each stage of the PRISMA flowchart are available from the authors upon reasonable request.

## Acknowledgments

DGK is funded by a post-doctoral award from Aston University College of Health and Life Sciences, awarded to JN & DGK.

## Notes

### Competing Interest Statement

The authors have declared no competing interest.

### Clinical Protocols

https://www.crd.york.ac.uk/prospero/display_record.php?ID=CRD42022343161

### Author Declarations

Source data is the records of publicly available research studies, openly available before the initiation of the research/systematic review. Records available via declared systematic review methodologies.

### Summary of Updates

Searches were rerun and results updated in March 2024, resulting in an additional 899 records screened and an additional 1 paper identified.

