## Supplemental Materials for "Limited Research Investigating the Value of MRI in Predicting Future Cognitive Morbidity in Survivors of Paediatric Brain Tumours: A Call to Action for Clinical Neuroimaging Researchers"

**Appendix 1 – Search Strategy**

**Web of Science**

| Block 1 Child  (TOPIC) | (pediatric OR paediatric OR infant* OR child* OR adolescen* OR youth OR teen* OR young OR neonat*) |
| --- | --- |
| Block 2 Brain  (TOPIC) | (brain OR CNS OR neuro* OR intracranial OR supratentorial OR cerebell* or “posterior fossa”) |
| Block 3 Tumour  (TOPIC) | (tumo$r OR neoplasm OR cancer* OR malignan* OR pilocytic-astrocytoma OR pituitary-tumo$r OR glioma OR neuronal-gial-tumo$r OR medulloblastoma OR ependym* OR astrocytoma OR craniopharyngioma OR germ-cell-tumo$r OR glioblastoma OR meningioma OR mesenchymal-tumo$r OR hemangioma OR atypical-teratoid*rhabdoid-tumor OR AT*RT) |
| Block 4 Cognition  (TOPIC) | (neuropsych* OR cogniti* OR “late-effects” OR mental OR IQ) |
| Block 5 MRI  (TOPIC) | (magnetic resonance imaging OR MRI OR DWI OR DTI OR diffusion*imaging OR ASL OR spectroscop* OR neuroimag* OR MRS OR perfusion OR MRSI OR GRE OR gradient-echo OR DSC OR DCE) |

**Search date:** 01/07/2022

**Number of results:** 3,449

**Search date:** 27/03/2024

**Number of results:** 77 (since previous search)

*TOPIC = title, abstract, author keywords, Keyword Plus

**Scopus**

| Block 1 Child  (TITLE-ABS-KEY) | (pediatric OR paediatric OR infant* OR child* OR adolescen* OR youth OR teen* OR young OR neonat*) |
| --- | --- |
| Block 2 Brain  (TITLE-ABS-KEY) | (brain OR CNS OR neuro* OR intracranial OR supratentorial OR cerebell* or “posterior fossa”) |
| Block 3 Tumour  (TITLE-ABS-KEY) | (tumour OR tumor OR neoplasm OR cancer* OR malignan* OR pilocytic-astrocytoma OR pituitary-tumour OR pituitary-tumor OR glioma OR neuronal-gial-tumour OR neuronal-gial-tumor OR medulloblastoma OR ependym* OR astrocytoma OR craniopharyngioma OR germ-cell-tumour OR germ-cell-tumor OR glioblastoma OR meningioma OR mesenchymal-tumour OR mesenchymal-tumour OR hemangioma OR atypical-teratoid*rhabdoid-tumor OR AT*RT) |
| Block 4 Cognition  (TITLE-ABS-KEY) | (neuropsych* OR cogniti* OR “late-effects” OR mental OR IQ) |
| Block 5 MRI  (TITLE-ABS-KEY) | (magnetic resonance imaging OR MRI OR DWI OR DTI OR diffusion*imaging OR ASL OR spectroscop* OR neuroimag* OR MRS OR perfusion OR MRSI OR GRE OR gradient-echo OR DSC OR DCE) |

**Search date:** 01/07/2022

**Number of results:** 2,941

**Search date:** 27/03/2024

**Number of results:** 503 (since previous search)

*TITLE-ABS-KEY = Title, Abstract, Keywords

**PsycINFO**

| Block 1 Child  (NOFT) | (pediatric OR paediatric OR infant* OR child* OR adolescen* OR youth OR teen* OR young OR neonat*) |
| --- | --- |
| Block 2 Brain  (NOFT) | (brain OR CNS OR neuro* OR intracranial OR supratentorial OR cerebell* or “posterior fossa”) |
| Block 3 Tumour  (NOFT) | (tumo?r OR neoplasm OR cancer* OR malignan* OR pilocytic-astrocytoma OR pituitary-tumo?r OR glioma OR neuronal-gial-tumo?r OR medulloblastoma OR ependym* OR astrocytoma OR craniopharyngioma OR germ-cell-tumo?r OR glioblastoma OR meningioma OR mesenchymal-tumo?r OR hemangioma OR atypical-teratoid*rhabdoid-tumor OR AT*RT) |
| Block 4 Cognition  (NOFT) | (neuropsych* OR cogniti* OR “late-effects” OR mental OR IQ) |
| Block 5 MRI  (NOFT) | (magnetic resonance imaging OR MRI OR DWI OR DTI OR diffusion*imaging OR ASL OR spectroscop* OR neuroimag* OR MRS OR perfusion OR MRSI OR GRE OR gradient-echo OR DSC OR DCE) |

**Search date:** 01/07/2022

**Number of results:** 811

**Search date:** 27/03/2024

**Number of results:** 69 (since previous search)

*NOFT = Anywhere except full text *(didn’t have option to limit to title, abstract and keywords)*

**Embase**

((pediatric OR paediatric OR infant* OR child* OR adolescen* OR youth OR teen* OR young OR neonat*) AND (brain OR CNS OR neuro* OR intracranial OR supratentorial OR cerebell* or “posterior fossa”) AND (tumo?r OR neoplasm OR cancer* OR malignan* OR pilocytic-astrocytoma OR pituitary-tumo?r OR glioma OR neuronal-gial-tumo?r OR medulloblastoma OR ependym* OR astrocytoma OR craniopharyngioma OR germ-cell-tumo?r OR glioblastoma OR meningioma OR mesenchymal-tumo?r OR hemangioma OR atypical-teratoid*rhabdoid-tumor OR AT*RT) AND (neuropsych* OR cogniti* OR “late-effects” OR mental OR IQ) AND (magnetic resonance imaging OR MRI OR DWI OR DTI OR diffusion*imaging OR ASL OR spectroscop* OR neuroimag* OR MRS OR perfusion OR MRSI OR GRE OR gradient-echo OR DSC OR DCE)).ti,ab,kw

**Search date:** 01/07/2022

**Number of results:** 1,028

**Search date:** 27/03/2024

**Number of results:** 177 (since previous search)

*ti,ab,kw = Title, Abstract, Keywords

IMPORTANT: If you copy-paste to rerun the search, remember to remove/retype quotation marks around “posterior fossa” and “late-effects” in search engine as it recognises these as illegal characters otherwise.

**MEDLINE**

((pediatric OR paediatric OR infant* OR child* OR adolescen* OR youth OR teen* OR young OR neonat*) AND (brain OR CNS OR neuro* OR intracranial OR supratentorial OR cerebell* or “posterior fossa”) AND (tumo?r OR neoplasm OR cancer* OR malignan* OR pilocytic-astrocytoma OR pituitary-tumo?r OR glioma OR neuronal-gial-tumo?r OR medulloblastoma OR ependym* OR astrocytoma OR craniopharyngioma OR germ-cell-tumo?r OR glioblastoma OR meningioma OR mesenchymal-tumo?r OR hemangioma OR atypical-teratoid*rhabdoid-tumor OR AT*RT) AND (neuropsych* OR cogniti* OR “late-effects” OR mental OR IQ) AND (magnetic resonance imaging OR MRI OR DWI OR DTI OR diffusion*imaging OR ASL OR spectroscop* OR neuroimag* OR MRS OR perfusion OR MRSI OR GRE OR gradient-echo OR DSC OR DCE)).ti,ab,kw

**Search date:** 01/07/2022

**Number of results:** 403

**Search date:** 27/03/2024

**Number of results:** 73 (since previous search)

*ti,ab,kw = Title, Abstract, Keywords

IMPORTANT: If you copy-paste to rerun the search, remember to remove/retype quotation marks around “posterior fossa” and “late-effects” in search engine as it recognises these as illegal characters otherwise.

**OSF Preprints (grey literature)**

((pediatric OR paediatric OR infant* OR child* OR adolescen* OR youth OR teen* OR young OR neonat*) AND (brain OR CNS OR neuro* OR intracranial OR supratentorial OR cerebell* or “posterior fossa”) AND (tumo?r OR neoplasm OR cancer* OR malignan* OR pilocytic-astrocytoma OR pituitary-tumo?r OR glioma OR neuronal-gial-tumo?r OR medulloblastoma OR ependym* OR astrocytoma OR craniopharyngioma OR germ-cell-tumo?r OR glioblastoma OR meningioma OR mesenchymal-tumo?r OR hemangioma OR atypical-teratoid*rhabdoid-tumor OR AT*RT) AND (neuropsych* OR cogniti* OR “late-effects” OR mental OR IQ) AND (magnetic resonance imaging OR MRI OR DWI OR DTI OR diffusion*imaging OR ASL OR spectroscop* OR neuroimag* OR MRS OR perfusion OR MRSI OR GRE OR gradient-echo OR DSC OR DCE))

**Search date:** 03/07/2022

**Number of results:** 1

Figure S1. PRISMA flow diagram from Original July 2022 searches

**Identification of studies via other methods**

**Identification of studies via databases and registers**

Records identified from:

OSF preprints (n = 1)

Literature reviews (n = 3)

Citation/reference searching (n = 4)

Records removed *before screening*:

Duplicate records removed
(n = 2,402)

Records marked as ineligible or unable to retrieve (n = 551)

Records identified from:

Web of Science (n = 3,449)

Scopus: (n = 2,941)

Embase: (n = 1,028)

PsycINFO: (n = 811)

MEDLINE: (n = 403)

**Identification**

Records screened

(n = 5,679)

Records excluded

(n = 5,517)

Reports not retrieved

(n = 0)

Reports sought for retrieval

(n = 8)

Reports sought for retrieval

(n = 162)

Reports not retrieved

(n = 9)

**Screening**

Reports excluded:

Age at diagnosis >18 (n = 52)

Neuropsych assessment precedes MRI (n = 52)

Not peer reviewed (n = 29)

No analyses of interest
(n = 10)

No standardised neuropsych assessment (n = 4)

Case study(s) (n = 1)

Non-English text (n = 1)

Reports assessed for eligibility

(n = 8)

Reports excluded:

Age at diagnosis >18 (n = 3)

MRI precedes neuropsych test (n = 3)

No analyses of interest
(n = 1)

Literature review (n = 1)

Reports assessed for eligibility

(n = 153)

Studies included in review

(n = 4)

**Included**
